# Semantic modelling of Common Data Elements for Rare Disease registries, and a prototype workflow for their deployment over registry data

**DOI:** 10.1101/2021.07.27.21261169

**Authors:** Rajaram Kaliyaperumal, Mark D. Wilkinson, Pablo Alarcón Moreno, Nirupama Benis, Ronald Cornet, Bruna dos Santos Vieira, Michel Dumontier, César Henrique Bernabé, Annika Jacobsen, Clémence M. A. Le Cornec, Mario Prieto Godoy, Núria Queralt-Rosinach, Leo J Schultze Kool, Morris A. Swertz, Philip van Damme, K. Joeri van der Velde, Nawel van Lin, Shuxin Zhang, Marco Roos

## Abstract

**Background:** The European Platform on Rare Disease Registration (EU RD Platform) aims to address the fragmentation of European rare disease (RD) patient data, scattered among hundreds of independent and non-coordinating registries, by establishing standards for integration and interoperability. The first practical output of this effort was a set of 16 Common Data Elements (CDEs) that should be implemented by all RD registries. Interoperability, however, requires decisions beyond data elements - including data models, formats, and semantics. Within the European Joint Programme on Rare Disease (EJP RD), we aim to further the goals of the EU RD Platform by generating reusable RD semantic model templates that follow the FAIR (Findable, Accessible, Interoperable, and Reusable) Data Principles.

**Results:** Through a team-based iterative approach, we created semantically grounded models to represent each of the CDEs, using the SemanticScience Integrated Ontology (SIO) as the core framework for representing the entities and their relationships. Within that framework, we mapped the concepts represented in the CDEs, and their possible values, into domain ontologies such as the Orphanet Rare Disease Ontology, Human Phenotype Ontology and National Cancer Institute Thesaurus. Finally, we created an exemplar, reusable ETL pipeline that we will be deploying over these non-coordinating data repositories to assist them in creating model-compliant FAIR data without requiring site-specific coding nor expertise in Linked Data or FAIR.

**Conclusions:** Within the EJP RD project, we determined that creating reusable, expert-designed templates reduced or eliminated the requirement for our participating biomedical domain experts and rare disease data hosts to understand description logic semantics. This enabled them to publish highly expressive FAIR data using tools and approaches that were already familiar to them.

## Background

The FAIR Principles [1] aim to provide guidance that will lead to an internet of data and services that is highly descriptive and machine-accessible, resulting in more extensive data discovery and reuse. FAIR data requires unambiguously identified entities to be richly described by unambiguously defined and identified concepts from thesauri and ontologies that are widely shared within a community and machine readable. When this is achieved, it will become much more straightforward to discover task-relevant data over distributed sites, accurately integrate those data, or analyse them by ‘data visiting’.

A significant barrier to Rare Disease research is that RD data is (a) extremely scarce, and (b) spread over many “boutique” repositories, often single-disease-specific and often curated by biomedically-oriented experts, who may not have access to experts in data or knowledge representation, capture or archival. In an initial step to address this, the EU RD Platform has begun to establish standards for integration and interoperability. The first practical output of this effort was a set of 16 Common Data Elements that should be implemented by all RD registries [2]. These include facets such as “sex”, “date of birth”, “age of onset”, and “diagnosis”, often together with a constraint on the allowed values of each of these data elements (for example, the possible values of ‘age at onset’ are ‘Antenatal’, ‘At birth’, ‘Date (dd/mm/yyyy)’, or ‘Undetermined’. Achieving uniformity of these 16 data facets, over all RD registries and biobanks, would be an excellent first-step towards enhanced discovery and reuse of these precious data.

Web-scale (which implies “mechanized”) interoperability, however, requires decisions beyond just a list of data elements - including data models, formats, and semantics. Within the European Joint Programme on Rare Disease, there is an aim to further the goals of the EU RD Platform by generating a “virtual platform” for interoperability between RD data assets - several hundred independent and non-coordinating data repositories - throughout Europe (and beyond). In part, this is being pursued by generating RD data that follows the FAIR Data Principles [3,4]. Historically, within this community, there have been efforts to train individual data owners to create FAIR data at-source. These have taken the form of annually recurring “Bring Your Own Data” [5] workshops (BYODs) where data owners meet FAIR experts and get hands-on experience in making their resources FAIR.

Because of their open-ended, exploratory structure, these BYOD events did not converge on a unified model for RD data, nor even the elements that should be included in those models. As such, the workshops primarily succeeded in raising awareness of FAIR, and the utility and benefits of following the FAIR Principles; however, the degree of inter-repository harmonization achieved by these workshops was extremely limited. Nevertheless, some preliminary data models were created at BYOD workshops, including the early version (V0.1.0) of CDE semantic model that is the focus of this manuscript.

In the case of the EJP RD project, it was immediately obvious that training individual participants in FAIR data modelling themselves was a non-starter - RD registries are limited by funding, expertise, and time. All three of those barriers make it infeasible for the “FAIRification” pathway of EJP RD to involve significant decision-making by the resource owner. Rather, we decided to centralize many of the decisions, ensuring that they were made by a small group of experts, and then disseminated outward to the individual participating registries and biobanks via a layer of registry liaisons who would communicate the needs, in both directions, between the data modelers and the registry custodians.

The final problem was how to enact the FAIRification itself - that is, how to do the “extract” and “transform” portions of the traditional ETL (Extract-Transform-Load) pipeline over resources that had no coordinating structure, and potentially no ability to code data transformation software themselves. Thus, we needed to create an ETL pipeline that could be deployed anywhere, over any native data structure, in highly secure privacy-sensitive environments, and execute a successful transformation using only the expertise that could be expected of most repository curators.

Here we describe the process of data modelling within the EJP RD, as applied to the set of CDEs defined by the EU RD Platform. We describe the semantic basis of those models, and how they have already been applied to distinctly different data types, showing that they have not been overly “fitted” to the data elements defined by the CDEs. Finally, we describe our current attempts to build an ETL pipeline that can fill these models, using a simple, structured Comma-Separated Value (CSV) export of source data from the originating registry hosts.

## Methods

### Modelling Activity

Modelling activities were undertaken via weekly meetings of a core group of EJP RD researchers with extensive experience in ontologies, knowledge representation, Linked Data modelling, and FAIR data. Meetings were carried out via Microsoft Teams meetings, where the model under discussion was presented via screen sharing.

As noted above, the European Platform on Rare Disease Registration has determined a set of 16 CDEs for RD registration. These are detailed in Table 1.

**Table 1:**
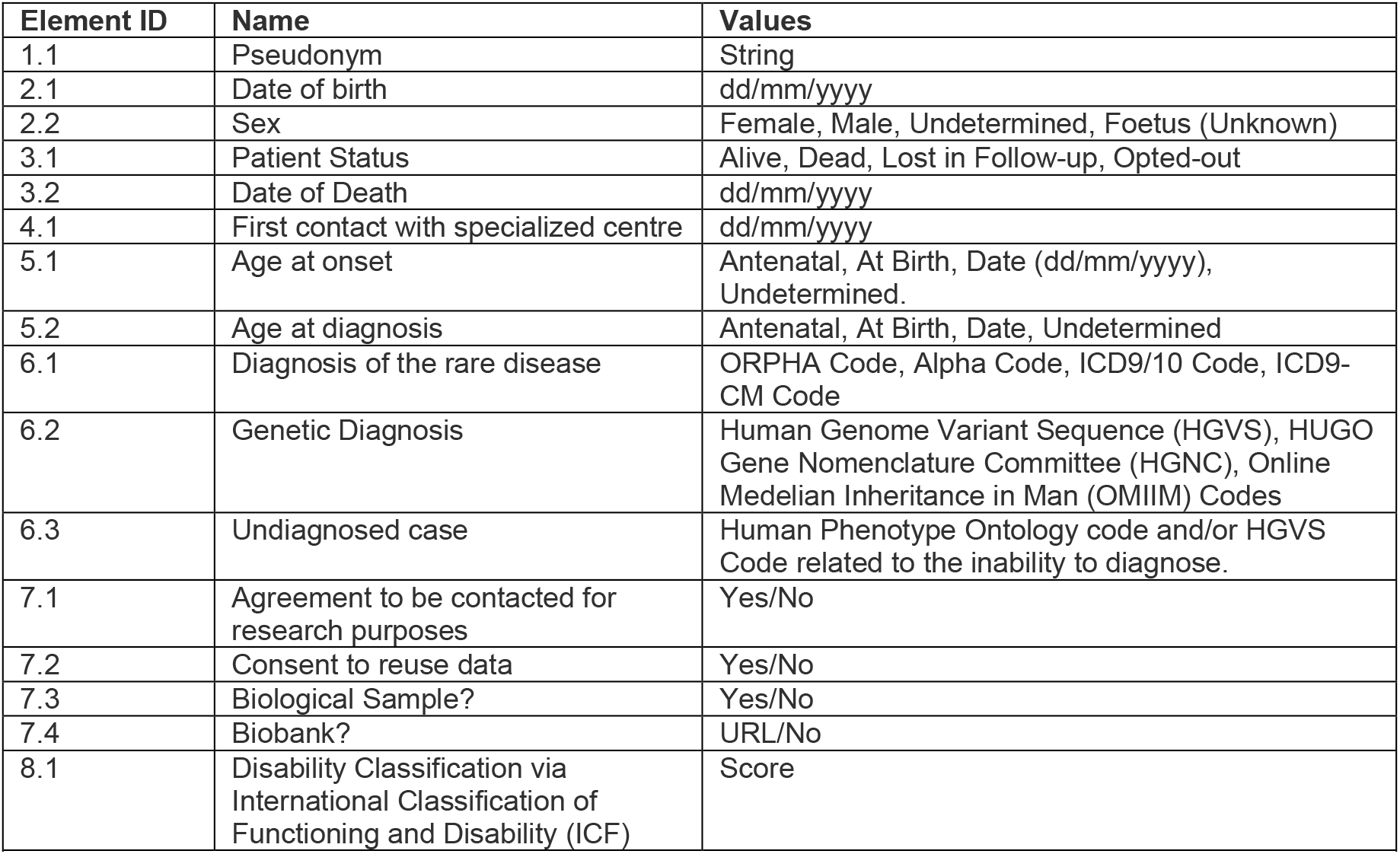
The European Platform for Rare Disease Registration set of Common Data Elements that should be made available by all rare disease registries.

Using these elements as a guide, together with additional documentation detailing how these elements should be filled, a first pass modelling phase [6] was undertaken where Linked Data representations for each CDE were constructed, using existing ontological terms or other shared Globally Unique Identifiers (GUID) wherever possible to model, for example, genotypes (Online Mendelian Inheritance in Man - OMIM Codes [7]) and phenotypes (Human Phenotype Ontology Codes [8]).

These first-pass models were then used to frame a more conceptual modelling process, looking at (for example) the inter-dependencies between the CDEs, the “nature” of the data – for example, is it obtained by questionnaire or by physical examination? - and what additional annotation would be useful to contextualize the CDE for correct interpretation (e.g., the date of the examination). There were several over-arching guidelines that constrained this modelling process:

1. We should use the minimum number of ontologies possible
2. We must strictly adhere to the ontological definition of a concept
3. The ontologies/vocabularies used must not have a restrictive license
4. The model should be designed in a forward-looking manner, anticipating other likely data elements, to minimize the need for future disruptive changes

Examination of the CDEs revealed that there were, in fact, many inter-dependencies between them – meaning that one CDE could not reliably be understood or contextualized without one or more of the others. For example, beyond CDE 1.1 - the patient pseudonym – all other CDEs depend on that one for their interpretation. For example, 4.1 ‘First contact with a specialized centre’ cannot be interpreted without a reference to the patient that made contact with the centre (via their pseudonym). Similarly, since individuals may have multiple diseases, each with its own diagnosis (CDE 6.1), the “age at diagnosis” (CDE 5.2) must somehow relate to the disease which was diagnosed at that age. In addition, we noted that most CDEs focused on data that would result from a formal interaction in a clinical setting, but those data gathering processes might be undertaken at different locations and times. For example, obtaining a biological specimen (CDE 7.3) would often be a surgical process, which would be undertaken in entirely different circumstances than the administration of a questionnaire to generate a disability score (CDE 8.1). Although the CDE requirements from the EU RD Platform do not require that this metadata be captured, it is nevertheless true that these details likely are being captured in many cases, thus it is useful to plan a model that can carry this contextual metadata, now or in the future. This minimizes the degree to which EJP RD participants would have to change their workflows to adapt to future changes.

We examined two ontologies that are commonly used in the life sciences to model processes and actors. The Provenance ontology (PROV-O) [9] aims at supporting representation of actions that agents undertake to generate or manipulate entities. The SemanticScience Integrated Ontology (SIO) [10] comprises elements of an upper ontology of types capable of modelling entities, processes, and qualities/attributes, together with relations that lead to a rigorous design pattern for associating these to one another. Conceptually, it is similar to Observ-OM [11]. SIO also has domain extensions, including biology and bioinformatics, that can help ensure that many clinical or biological concepts are being used in a logically sound manner.

We selected SIO for a number of reasons. For example, SIO has the capacity to represent data content – that is, while PROV-O has the concept of an Entity, which could represent the output of a process, it does not have the other properties or concepts that can describe what that entity is, or its value, or units. Conversely, SIO includes CDE-relevant concepts such as “medical diagnosis”, which allows us to use SIO-defined properties and classes for the majority of the core CDE model. Similarly, PROV-O has no way of representing attribute/quality relations of an individual. Given that almost all of the CDEs are measurements of some attribute of the patient who participated in a clinical process, the inability to associate the output of a process (like a phenotype) as an attribute of the participating patient would make a PROV-O model highly unintuitive for our target end-users. As such, while PROV-O might be useful at a later date to describe, for example, the precise details of a Phenotyping or Genetic Diagnosis workflow, our needs in this modelling exercise are distinct, and are better represented by the SIO upper-level concepts.

Following the documented design patterns for SIO [12] we derived the core model shown in Figure 1. Some of the rationale for this model are as follows: First, all CDE observations are, in some way “about” an individual patient. As such, it is necessary to connect patients to these observations. In some cases, the CDEs pertain to a direct attribute of the patient (e.g., their birth date). In other cases, the CDE is not an attribute of the patient *per se*, but rather the connection between patient and the CDE is via an action or activity that the patient engaged in; for example, the first interaction of a patient with a rare disease expert centre. Certainly, for all CDEs there is at least the process of recording the information, and as such, we decided that a “process” was a concept shared by all CDEs. Early discussions also raised the issue of an individual having multiple roles in the healthcare system, for example, being both a patient and a physician. As such, it was necessary to connect a participant to the process indirectly, by declaring the role they play as the process becomes realized. An individual may have many roles, and we determined that in every case, there was a distinct identifier that was assigned to that role – for example, a driver’s license ID is assigned to one’s role as a driver, and a student ID is assigned to one’s role as a student, yet both identifiers may apply to the same individual. Finally, processes have outputs, where those outputs (often) refer to some measurement of an attribute of the patient. The attribute, and its measurement, are distinct – for example, all patients share the attribute of “sex” but for some patients this attribute has the value “male” and for others it has the value “female”. Combining these considerations leads to the core model shown in Figure 1, where there are 5 “kinds” of things: entities (individuals, and measurements), roles, processes, attributes, and identifiers. While there are additional relationships between these concepts, we removed all but the relations required to connect the model. This will simplify the creation of query systems, by limiting the possible ways the model can be explored, better enabling the construction of reusable query templates.

**Figure 1:**
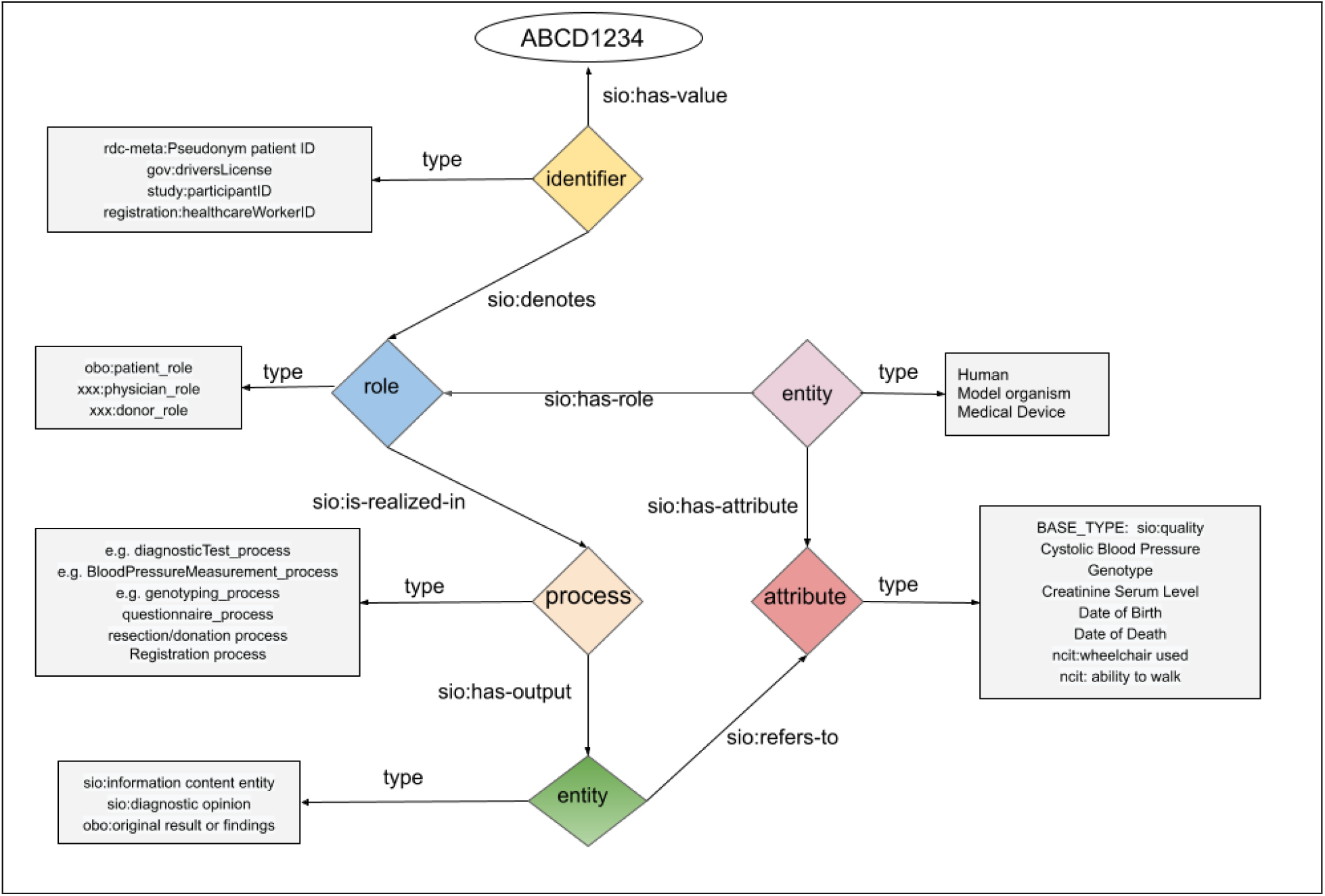
Conceptual diagram of the overall SIO model to be applied to the CDEs. It is centred around 5 primary elements – identifiers, entities (physical and information-content), roles, processes, and attributes. In the diagram, we provide hypothetical examples of the specific ontological types that might be associated with each element.

Using this high-level model as a guide, the EJP RD semantic modelling group then reiterated the process of examining each CDE and, through Teams meetings and dedicated “designathons” we reached agreement on which portions of the high-level model were appropriate for each CDE, and what the ontological type constraints (square boxes in Figure 1) should be for the elements of that specific CDE model. Through this process, we also identified what we refer to as a “base type” for each of the elements, for example, the process node is always typed as “sio:process”. In this way, if there is not a more specific type assigned to the model node, we still maintain the best practice of having all nodes in our model ontologically typed. These “base types” are built into our transformation templates (described below) and require no knowledge by the end-user.

### Extract-Transform-Load

Registries participating in the EJP RD have a wide range of underlying infrastructures and data management and curation expertise, ranging from well-established commercial enterprises such as Castor [13], open source data platforms such as MOLGENIS [14,15] to smaller, parent-run organizations even using spreadsheets to capture data. Therefore, the task of finding a “lowest common denominator” for a transformation workflow that can be executed by custodians at any level of expertise was a high priority. Moreover, a primary objective of EJP RD is to encourage FAIRness, thus the solution itself should be reusable.

Through discussions with EJP RD partners, it became clear that there was a preference for very straightforward data structures such as CSV, since this is an exchange format that can be derived easily from any of the more complex formats. As such, we identified RDF Mapping Language (RML) as a potential target technology, as it is capable of modelling reusable templates that support CSV to RDF transformations. The creation of these CSV files is left as an exercise for each registry owner, as their individual situations will be diverse. Considerations for the registry owners include:

- Ensuring date formatting is correct (ISO 8601)
- Ensuring that any abbreviated ontology terms have been converted into their equivalent full URLs (e.g., Human Phenotype Ontology terms must be represented by their URL in the CSV file)
- Ensuring any data elements have been modified to match the documented constraints (e.g., conversion of textual descriptors into ontology term URIs)
- Ensuring that every row is uniquely identified (for this purpose, we have established a web service that can be called from MS Excel or a custom script that generates a unique identifier based on a timestamp, since MS Excel has no inherent capability to generate GUIDs without custom coding)

With the aim of simplifying the somewhat complex RML syntax, such that our EJP RD FAIRification stewards, or potentially the registry data stewards themselves, could edit the template if required, we identified a second, related technology – YARRRML [16] – which is a human-readable way to declare RML transformation rules, using YAML (YAML Ain’t Markup Language) as the syntax. YARRRML documents can be converted into RML templates, which can then be automatically applied to CSV files to achieve their transformation.

The transformation itself is done by “RDFizing” software. We tried two packages – RMLMapper [17], and SDM-RDFizer [18]. RMLMapper has a rich set of features, including the ability to encode transformation rules that can trigger execution of algorithms over a CSV cell prior to the RDF transformation. SDM-RDFizer conversely, lacks these powerful extensions, but is significantly faster in our (informal) head-to-head tests. Since YARRRML currently cannot encode rules, we do not benefit from the additional power provided by RMLMapper, and thus selected SDM-RDFizer for this modelling initiative. Nevertheless, the choice of RDFizing technology can be revisited at a later date, without affecting any of our other decisions.

For storage of the resulting Linked Data, we have selected GraphDB [19], due to its ongoing support by the developers, the availability of a free (though not open source) version, and the availability of a fairly comprehensive API for mechanization of data loading, maintenance, and querying. GraphDB also supports different access control methods which provides options for securing access to the FAIRified dataset. A “bootstrapping” docker image for GraphDB was created to ensure that GraphDB is installed and configured correctly, thus eliminating the need for the registry host to have this expertise.

Deployment of the ETL pipeline is achieved via docker-compose, where every component is packaged as a docker image and uses a docker [20] network to facilitate communication between the components. This ensures that there are no unnecessary ports or APIs exposed on the registry server, helping maintain the security of their internal space. The three components mentioned above - RMLMapper, SDM-RDFizer, and GraphDB - are coordinated via a fourth docker container, representing an orchestration tool. The orchestrator is activated by a Web call to its interface. Once initiated, it automatically refreshes the current database of YARRRML templates from the EJP RD GitHub, and then examines the content of a folder shared with the host. This shared folder contains the host’s CSV files that will be subject to RDF transformation. Using filename-matching, the system matches each CSV with an appropriate YARRRML template and executes the transformation. After all transformations have completed, a connection is opened to GraphDB, all previous data is deleted, and the refreshed data is uploaded.

The suite of four docker images are referred-to as the “CDE-in-a-Box”, and the instructions for running the bootstrapping process, as well as how to interact with CDE-in-a-Box, are available on a dedicated Git [21].

### Testing

Speed tests were run by calling RMLMapper and SDM-RDFizer images via docker-compose locally. A variety of exemplar 10.000 row CSV files and YARRRML templates were used for the measurement and execution process. The average speed of RDF triple generation was 12500 triples per second. The tests were run on an AMD Ryzen 7 3800XT 3.9 GHz CPU workstation, with 32 Gb 3200Mhz RAM memory, RTX 2070 Super 8 Gb GPU and M.2. NVMe SSD memory. Quality-control tests will, largely, be registry-specific, though we are considering possible mechanisms for generalizing this problem also.

## Results

### The Models

The models created to capture the 16 CDEs are described in Table 2, and are available in the project GitHub [22].

**Table 2.** Models created to represent the CDEs. Models are created in YARRRML and made available on the project GitHub, accompanied by markdown documentation explaining the structure of an appropriate CSV file. Note that not all EU RD CDEs appear 1-to-1 with a CDE model. This is because, for example, the consent CDE can be reused for diverse types of consent (e.g., consent for contact, consent for data reuse), and the Pseudonym CDE is a part of every other model, and therefore has not been modelled as an independent element.

| CDE Model Name | Purpose |
| --- | --- |
| Disease Progression: [23] | A “container” node to group together all other CDEs that refer to the same diagnosis. For example, the “age of diagnosis” CDE is related to a specific rare disease via traversal into the “disease progression” container, and then traversal into the “diagnosis” CDE that is also connected to “disease progression” |
| Care Pathway [24] | Captures the date of first contact with the specialist healthcare system; is connected to “disease progression” |
| Diagnosis [25] | Captures the final disease diagnosis using ORPHA codes; is connected to “disease progression” |
| Disease History [26] | Captures age at first symptoms and age at diagnosis; is connected to “disease progression” |
| Genetic Diagnosis [27] | Captures the sequence variant(s) found in this patient, using a variety of different coding systems; is connected to “disease progression” |
| Patient Consent [28] | Captures the consent of the patient over several axes (e.g., consent for contact, consent for data reuse). Provides a reference to the signed consent form, as well as an input reference to the (blank) consent template. |
| Patent Status [29] | Captures the current status of the patient, and their date of death if the patient is deceased |
| Personal Information [30] | Captures (superficial) personal information such as birth date and sex (there are ongoing debates in the EJP modelling group as to whether this should be converted to an age, or an age-range, for improved privacy) |
| Phenotyping [31] | Captures the phenotypes of the patient, using Human Phenotype Ontology terms |
| Disability [32] | Captures the score for a disability test. The specific test administered is indicated as one of the child nodes of obo:NCIT_C20993 (Clinical or Research Assessment Tool), and thus this CDE model is broadly useful for many disorders. |
| Undiagnosed [33] | Captures the case where a patient has phenotypic anomalies, and an identified sequence variant, but for some reason has not been definitively diagnosed. |

To help registry owners and data stewards understand the models, they are generated and published in a variety of formats. The YARRRML is accompanied by an exemplar “runnable” CSV file, which is in turn documented by a Markdown document containing a description of the CDE Model, its intended use, the CSV column headers, the constraints on the content of each column, and any usage notes that will assist the data owners in their understanding of how to generate compatible CSV. A screenshot of the documentation is provided in Figure 2.

**Figure 2.**
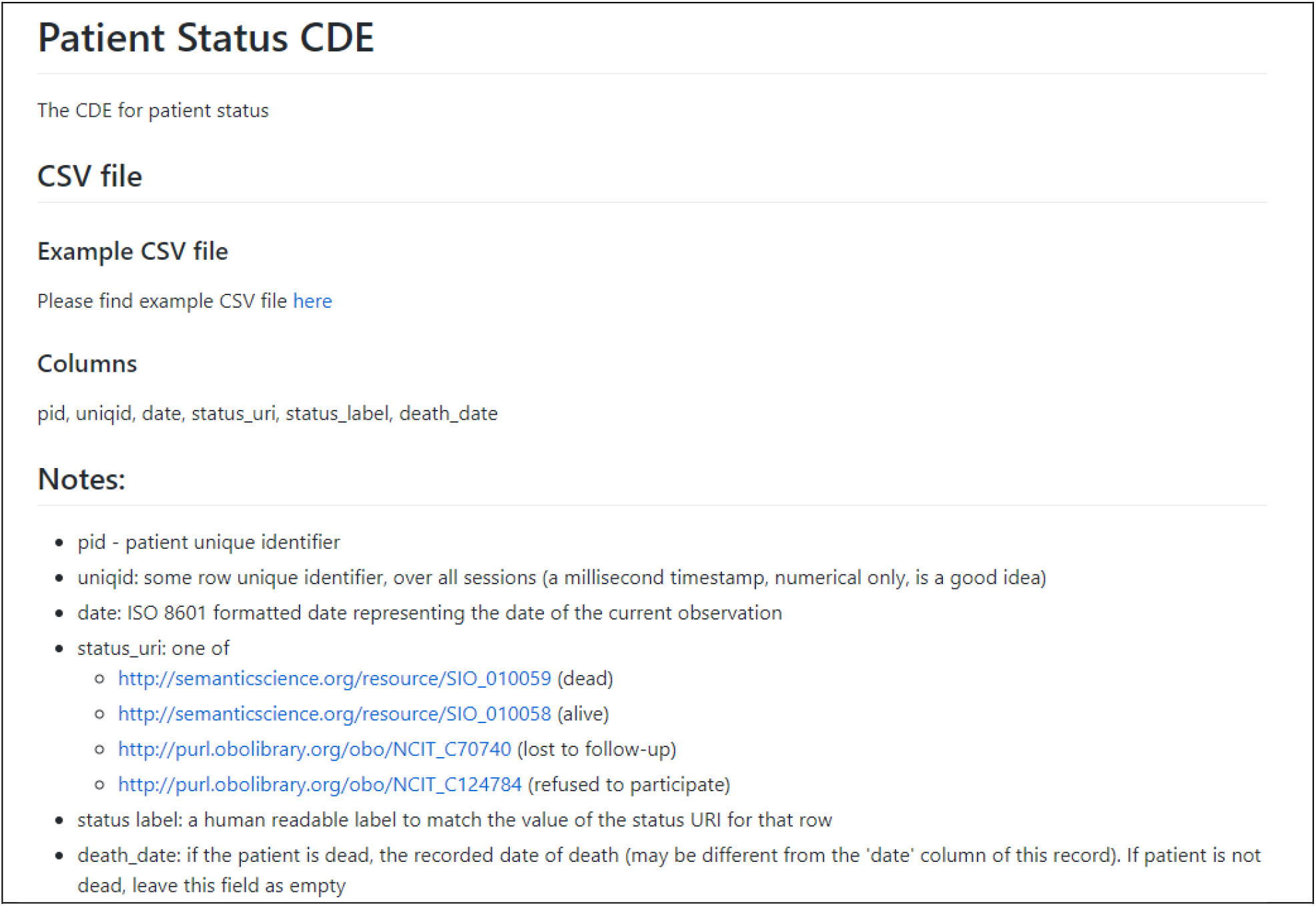
The Markdown documentation explaining how to prepare a CSV file for the “Patient Status” CDE. Documentation includes, where appropriate, the restrictions on the possible values in a given column, such as ‘status uri’ in this example.

To assist both data owners and data consumers, a variety of other representations are also generated. When the exemplar CSV is run through the transformation pipeline, the resulting RDF file is then converted into a model image via a semi-automated mechanism [34]. A Shape Expression (ShEx) model is also created to allow data owners (and users) to validate these transformations. The ShEx models are manually created according to 2.1 primer specification [35], and the resulting ShEx file is converted into an image via the RDFShape tool [36]. An exemplar RDF visualization for CDE #3 “Patient Status” is diagrammed in Figure 3, and a diagram of the ShEx validator for that model is shown in Figure 4.

**Figure 3.**
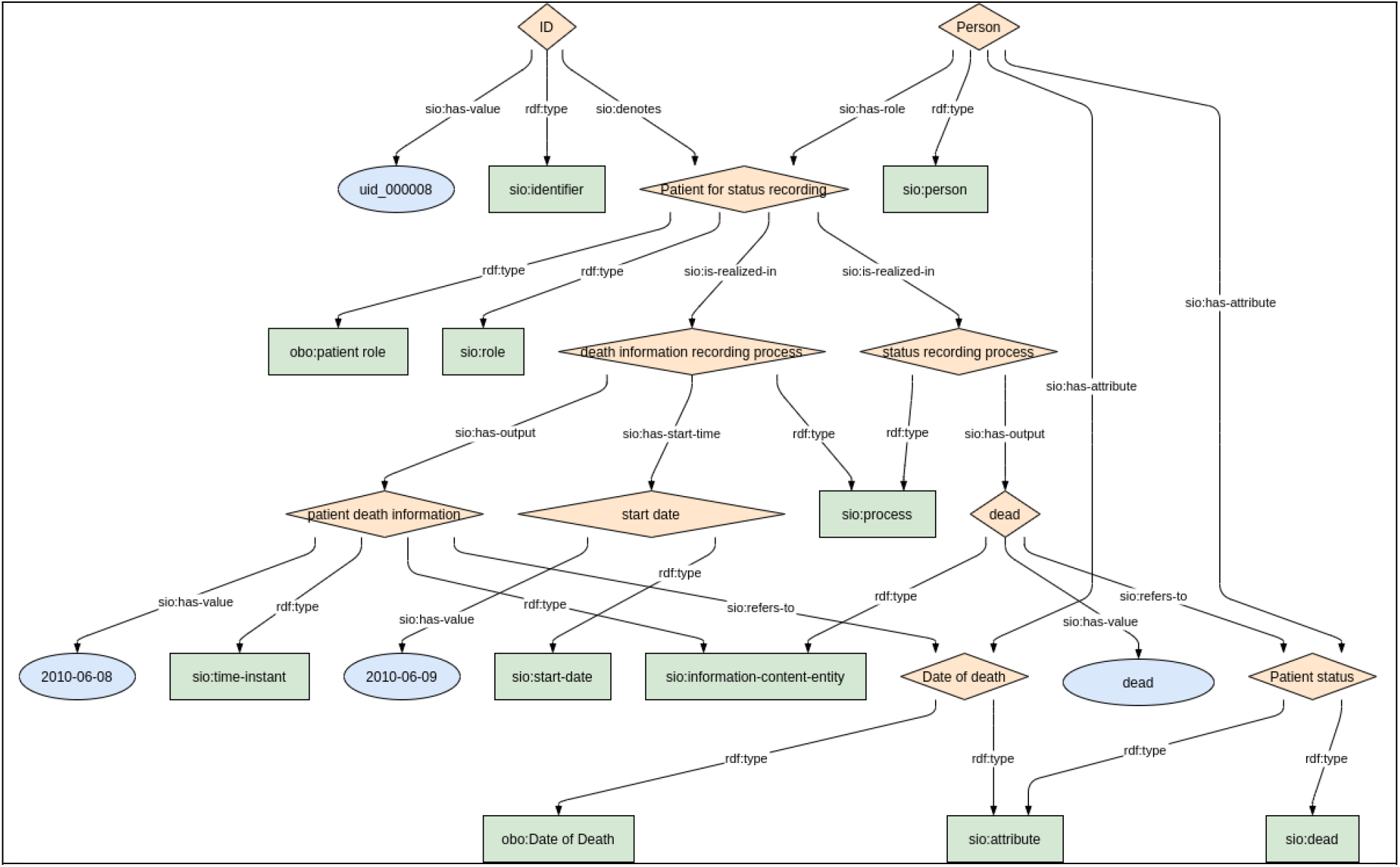
Visualization of an exemplar RDF instance for the “Patient Status” CDE (CDE 3.1 & 3.2).

**Figure 4.**
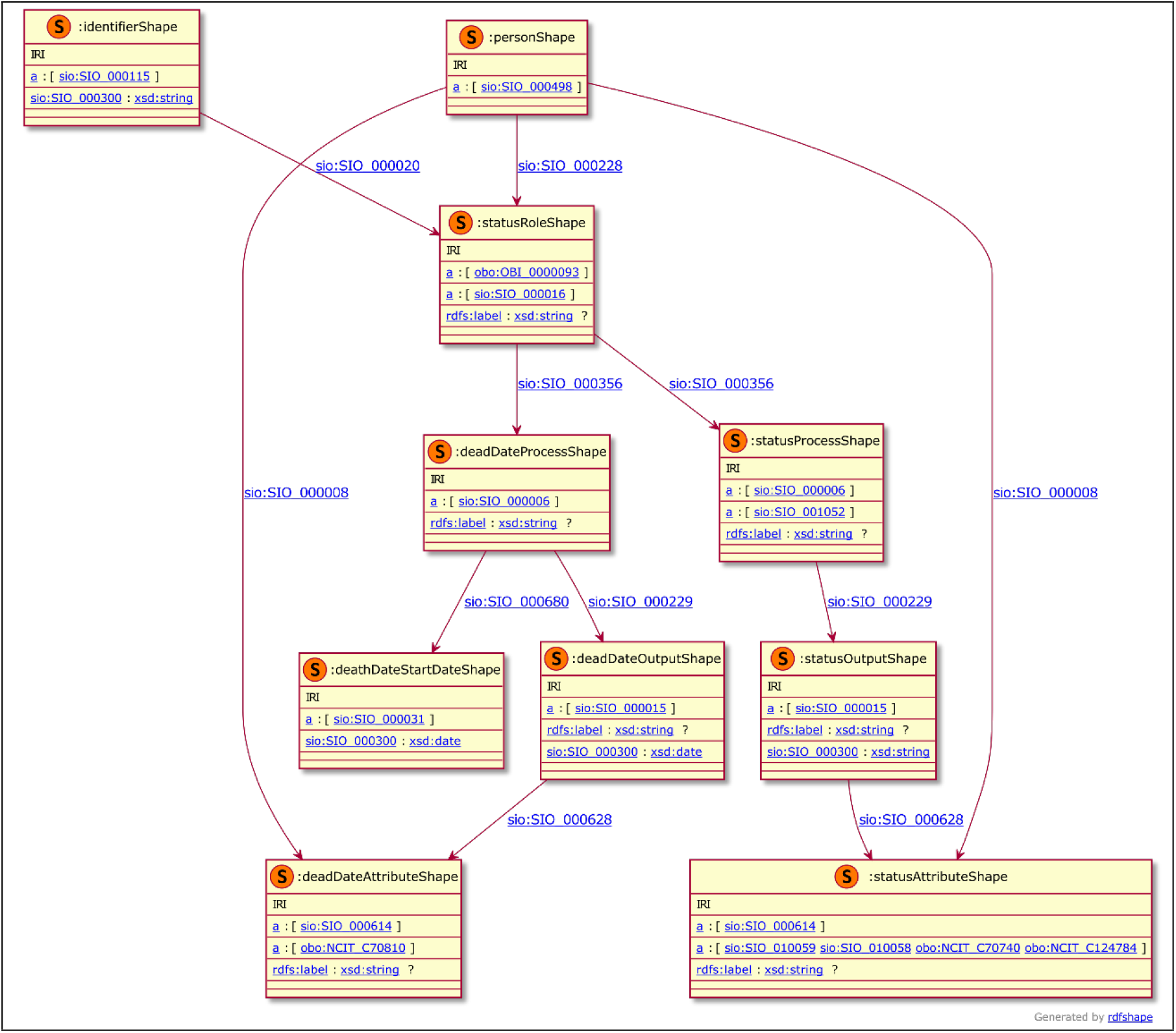
Visualization of the ShEx validation shape for the Patient Status CDE data.

### Model Filling - “CDE in a Box”

As described in the Methods section, the CDE-in-a-Box is deployed via docker-compose and is triggered by a simple HTTP GET on a localhost URL. The speed of the transformation using the default configuration of the current docker images is approximately 12,500 triples per second.

## Discussion

When undertaking any modelling activity, there is always the potential to “over-fit” the model. To this end, we have been attempting to apply the model to datatypes other than those covered in the CDE list. Specifically, we have looked at three very distinct datatypes: physical body measurements, laboratory tests, and Patient-Reported Outcome Measurements (PROMs), which are a questionnaire-style metric. In all cases, we were able to generate the Linked Data record with few or no changes to the core model. In particular, the Physical Body measurements required only an additional link to a measurement protocol; for PROMs we added an Input to the Process node representing the PROM question; and for Laboratory Tests we extended this further where an Input is included - constrained to being a body tissue - a “target” is included - constrained to being the compound being measured - and link is added to the measurement protocol document (see Figure 5). Hence, we believe that this core model is capable of representing the majority of data entities we will encounter in the biomedical/clinical space with only minor modifications.

**Figure 5.**
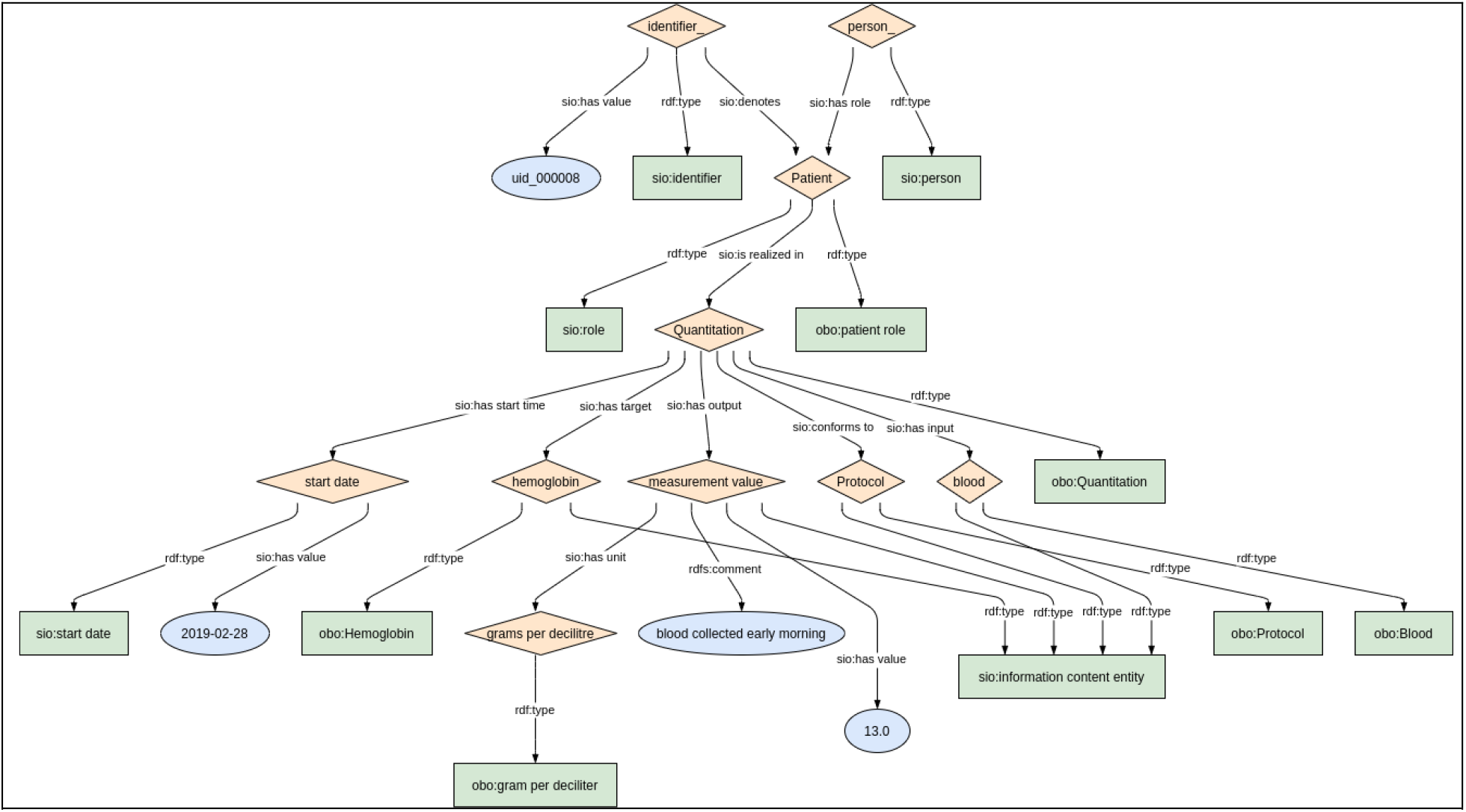
The model for Laboratory Measurements. Of note are the three new connections on the “Quantitation” (Process) node – one representing the input (blood), one representing the target molecule (haemoglobin), and the third representing the link to the protocol. The remainder of the model is (structurally) identical to the core model shown in Figure 1.

With respect to generalizability and scalability of this approach, a comprehensive survey of the European Reference Networks (ERNs) participating in EJP RD revealed 13 categories of data from 16 ERN data dictionaries; for example, “laboratory tests” and “personal information” are two such categories. Every category requires a YARRRML template to be constructed, following the core pattern but changing, for example, the default ontological types of each node, and the column header names. We have built code libraries that automatically generate these YARRRML templates via a simple API, and thus in practice, a new YARRRML model can be created in approximately an hour now that the general pattern has been established. Documentation of the model, and decisions about the constraints on the allowed content of each CSV column takes more consideration and time, though all elements of a well-documented model can be easily created in less than a day. This, however, leads to a problem for which the correct solution is, as yet, not known. Because the models themselves are generalized, the problem of selecting the correct specific value for a given column becomes a task for the data provider. For example, in the Body Measurement model, we document that the “attribute being measured” column should contain an ontology Uniform Resource Identifier (URI) that is a child of obo:NCIT_C19332 (personal attribute). While many of the participating hosts are familiar with ontologies, “coding” (the act of assigning a controlled vocabulary term to a concept, observation, or phenomenon) is an activity primarily undertaken by insurance and governmental organizations, and by trained disease classifiers, and as such many other participants will not have this experience. Thus, we suspect that this task may be difficult for a subset of our registry participants. One alternative is that the FAIR experts create a specific model for every case (every attribute, every lab measurement, etc.). This, however, would result in many highly specific models, and would in turn, require the data host to generate separate CSV files for each model. The alternative is to keep the models generic and find another way to provide advice or support to the data hosts as they generate the CSV. We are exploring both solutions to gain a better understanding of how to address this problem in the future.

Further, the transformation step itself – generating ~12,500 triples per second – is also sufficiently fast that it would be possible to generate a new snapshot on a nightly basis. Finally, the models are intended to be reusable, and the onus of creating a matching CSV is put on the data owners/experts. Another approach would have been to create a comprehensive transformation map of the entire dataset, for every participating registry using, for example, R2RML [37]. Since the data owners are non-experts, this approach would have centralized the problem of mapping into the hands of the few Linked Data experts in the EJP RD, which we feel is a much less practical solution, and less scalable. The solution proposed here distributes the effort over many more participants, and the sharing of a set of core models ensures that, despite being non-coordinating, the participating registries will nevertheless generate interoperable outputs.

Our plans are to build a quality-checking layer, using the ShEx to ensure that the output RDF from each site is conforming to the expected model. In addition, we will soon begin to provide training to those who wish to learn how to build the YARRRML themselves (either manually or using our code libraries) such that they can expand into other datatypes, or add new metadata facets (e.g., start and end times on processes) without necessarily requesting help from the EJP RD modelling team. In this way, we hope that the core data will be interoperable, even if individual sites add enhanced metadata that is not in-common with other registries.

## Conclusions

We undertook a process of constructing a reusable, generic data model, based on the design principles of the Semantic Science Integrated Ontology, to represent all the EU Rare Disease Platform Common Data Elements. Emergent mapping technologies such as YARRRML, RML, and “RDFizing” tools allowed us to create an automated pipeline for filling these data models starting from a well-documented CSV template – a format accessible to all our target end-users. We demonstrated the generic nature of the model by successfully extending it - while remaining within the overall architecture of SIO - to widely disparate non-CDE clinical data within the Rare Disease space. Feedback from end-users indicates that they found this solution helpful, and easy to apply. As FAIR data publishing becomes increasingly an expectation – even a requirement – of funding agencies and publishers, there is an urgent need for straightforward tooling to assist data providers to comply with these expectations. In many cases, those who generate data will not have expertise in data modelling, and particularly not in semantically grounded data modelling, as is a requirement of FAIR. The activities and workflows described here indicate that the approach of building a generic, reusable, models, and an automated pipeline to fill them, will be widely applicable in biomedicine and beyond.

## Data Availability

All data and code is available under open source licenses at the links provided in the manuscript text.

## List of Abbreviations

API: Application Programming Interface
BYODs: Bring Your Own Data workshops
CDE: Common Data Element
CSV: Comma Separated Values
EJP RD: European Joint Programme on Rare Disease
ERN: European Reference Network
ETL: Extract, Transform, Load
FAIR: Findable, Accessible, Interoperable, Reusable
GUID: Globally Unique Identifier
OMIM: Online Mendelian Inheritance in Man
OWL: Web Ontology Language
PROM: Patient Reported Outcome Measures
PROV-O: Provenance Ontology
RD: rare disease
RDF: Resource Description Framework
RML: RDF Mapping Language
ShEx: Shape Expression
SIO: SemanticScience Integrated Ontology
URI: Uniform Resource Identifier
YAML: YAML Ain’t Markup Language

## Declarations

### Ethics approval and consent to participate

Not applicable

### Consent for publication

Not applicable

### Availability of data and materials

Project name: CDE Semantic Model Implementations

Description: Code that generates the YARRRML, and the docker-compose repository used by “CDE in a Box”; sample CSV data and CSV documentation.

Project home page: https://github.com/EJPRD-vp/CDE-semantic-model-implementations

Operating system(s): Platform independent

Programming language: Ruby

License: MIT

Project name: CDE Semantic Model

Description: Diagrams, ShEx, and sample RDF for each of the Semantic Models

Project home page: https://github.com/ejp-rd-vp/CDE-semantic-model

Operating system(s): Platform independent

License: MIT

Project name: CDE in a Box

Description: Bootstrap and Production docker-compose files to run CDE in a Box

Project home page: https://github.com/ejp-rd-vp/cde-in-box

Operating system(s): Platform independent

License: Apache 2.0

### Authors’ contributions

All authors contributed to this work through participation in model design workshops, weekly discussions, and writing and revising this manuscript. Specific additional contributions are as follows: MDW & RK created CDE-in-a-box code and docker images; RK created workflow and code to generate RDF model images; MDW created YARRRML template files, sample CSV files, and markdown documentation; MD provided guidance on SIO modelling, expanded SIO design patterns and edited the SIO ontology; RC & MR cross-referenced CDE models with other clinical data models and checked all semantics for the modelling group; PA executed quality control of workflow outputs, creation and review of images for RDF and ShEx models; BdSV reviewed user instructions, updated documentation, tested CDE in a box Platform; NB planned and hosted “designathons” and other workshops, and gathered feedback; LJSK co-authored the first draft of the CDE model; PvD reviewed parts of the model for ontological consistency; NvL and the Duchenne Parent Project initiated the code development project that led to the CDE in a Box transformation pipeline, and provided end-user review and feedback; SZ reviewed RDF and ShEx files; BdSV, SZ, CHB, JKvdV, CLC executed surveys of registries and collected feedback from end-users.

## Acknowledgements

We would like to express our gratitude to the Duchenne Parent Project in the Netherlands for allowing the code for portions of their FAIR transformation solution to be open source and included in the CDE in a Box. This solution was specifically commissioned and developed for their patient-led registry - The Duchenne Data Platform - which had undergone a FAIRification process in 2021. Their generosity stems from their continuous belief in FAIR as a new paradigm for optimising data visiting and analysis and as a result, pledged to support others in their own FAIR data endeavours. We thank Foundation 29, the software developers of the Duchenne Data Platform, for their technical expertise and positive collaboration during the development of the Duchenne FAIR project. We furthermore wish to thank Leo Schultze Kool for helping us start the CDE modelling process for the FAIRification of the VASCA registry, his support for FAIR implementation of registries, and valuable feedback on the interpretation of CDEs from a clinical perspective, and Peter-Bram ‘t Hoen for his continuous active support of our efforts. Finally, the authors acknowledge the support of Ana Rath and Franz Schaefer in the development of a conceptual framework for the EJPRD Virtual Platform.

